# Genome-wide association study of skin and soft tissue infection susceptibility

**DOI:** 10.1101/2020.09.03.20187468

**Authors:** Tormod Rogne, Kristin V Liyanarachi, Humaira Rasheed, Laurent F Thomas, Helene M Flatby, Mari Løset, Dipender Gill, Stephen Burgess, Cristen J Willer, Kristian Hveem, Bjørn O Åsvold, Ben M Brumpton, Andrew T DeWan, Erik Solligård, Jan K Damås

## Abstract

**Background:** Skin and soft tissue infections (SSTIs) are common worldwide, but little is known about the genetic susceptibility and the causal effect of cardiometabolic risk factors. We therefore conducted the first genome-wide association study (GWAS) of SSTIs, with downstream analyses including Mendelian randomization analyses.

**Methods:** The GWAS was conducted using the UK Biobank as discovery cohort, with 6,107 cases and 399,239 controls, and the Trøndelag Health Study (HUNT) as replication cohort with 1,657 cases and 67,522 controls. Cases and controls were those who had or had not been hospitalized with an SSTI diagnosis, respectively.

**Findings:** One locus, lead single-nucleotide polymorphism rs3749748 in *LINC01184/SLC12A2*, was associated with SSTIs in the UK Biobank (odds ratio [OR] 1.19, p-value = 7.6e-16) and replicated in HUNT (OR 1.15, p-value = 6.3e-4). Meta-analysis confirmed the lead variant (OR 1.18, p-value = 4.4e-18), as well as suggested two additional loci close to genome-wide significance (rs2007361 in *PSMA1*, OR 0.91, p-value = 5.1e-8; and rs78625038 in *GAN*, OR 1.86, p-value = 5.9e-8). Gene-based association tests identified four genes linked to SSTIs: *SLC12A2, PSMA1, GAN*, and *IL6R*. The minor allele of rs3749748 reduced the gene expression of *SLC12A2* primarily in monocytes and macrophages. Mendelian randomization analyses showed that increasing body mass index and lifetime smoking habits increased risk of SSTIs.

**Interpretation:** *LINC0118/SLC12A2* was robustly associated with SSTI incidence and may exert its effect through reduced gene expression in monocytes and macrophages. Reducing tobacco smoking, overweight and obesity in the population may reduce the incidence of SSTIs.

## INTRODUCTION

Microbial invasion of the skin and underlying soft tissues, known as skin and soft tissue infections (SSTIs), contribute to considerable burden of disease worldwide.^1,2^ The vast majority of these infections are bacterial, and from Gram-positive bacteria such as streptococci and staphylococci in particular.^1^ Studies from the ambulatory and inpatient setting in the US have observed an incidence of roughly 50 SSTIs per 1,000 person-years, far greater than the incidence rates for urinary tract infections and pneumonias combined.^3,4^

Given the high disease burden of SSTIs it is important to identify host factors of the disease, novel therapeutic targets, and subjects at an increased risk. A range of risk factors - cardiometabolic in particular - are suspected to increase the risk of SSTIs, including obesity, type-2 diabetes, and smoking.^1,5,6^ While traditional epidemiological studies are subject to confounding and reverse causation, one may make use of the fact that genetic variants are randomly allocated at conception to construct pseudo-randomized studies, known as Mendelian randomization (MR) analyses.^7^ Increasing BMI has been found to increase the risk of SSTIs in such a framework,^5,6^ but other cardiometabolic risk factors have not been explored. The genetics of susceptibility to SSTIs remain largely unknown, and the only published genome-wide study on SSTIs to date is a family-based linkage study of 52 Finnish families that did not manage to identify significant linkage to any genes for erysipelas or cellulitis susceptibility.^8^

In order to evaluate the genetic susceptibility of SSTIs, we have conducted the first genome-wide association study (GWAS). Two independent European cohorts were used, in which a total of 7,764 subjects hospitalized with SSTIs served as cases and 466,761 subjects not hospitalized with SSTIs served as controls. To explore possible biological pathways and processes for the identified genes, we conducted downstream in-silico analyses, including gene expression analyses and linkage disequilibrium (LD) score regression. Finally, we performed MR analyses to identify potential causal relationships between cardiometabolic risk factors and SSTIs.

## METHODS

### Study populations

This study consisted of two independent cohorts, where the UK Biobank served as the discovery cohort in the genetic association analyses, while the the Trøndelag Health Study (HUNT) served as the replication cohort.

### UK Biobank

Details about the UK Biobank have previously been described.^9^ In brief, the cohort consists of 503,325 subjects enrolled between 2006 and 2010 throughout the United Kingdom. Age at baseline was between 38 and 73 years, and 94% were of self-reported European ancestry. At baseline, genome-wide genotyping was done on 488,377 individuals, including 84% of self-reported white-British ancestry with European genetic ethnicity. Information on self-reported health and lifestyle was collected, along with measurements such as height and weight. Inpatient hospital data on all participants was available through electronic record linkage.

### HUNT

The HUNT Study is a series of surveys conducted in the Nord-Trøndelag region in Norway (∼130,000 inhabitants) between 1984 and 2019 on subjects 20 years and older.^10^ We used data from HUNT2 (1995–1997) and HUNT3 (2006–2008), in which 78,973 subjects representative of the adult Norwegian population participated.^10^ Baseline characteristics were collected at study enrollment, and selected measurements were made including height and weight. Information on all hospitalizations in the county and to the regional tertiary care hospital were linked to the study subjects. Through linkage with the Norwegian population registry, we retrieved data on date of emigration out of the study region and date of death.

### Phenotype

Cases and controls were defined the same way in UK Biobank and HUNT. The following International Classification of Diseases (ICD)-9 and ICD-10 codes were considered as SSTI codes: 035 (erysipelas; ICD-9), 729.4 (fasciitis, unspecified; ICD-9), A46 (erysipelas; ICD-10), L03 (cellulitis and acute lymphangitis; ICD-10), and M72.6 (necrotizing fasciitis; ICD-10). These codes are used primarily for bacterial infections, and non-bacterial infections of the skin have other specific codes not considered. In our main definition of SSTI, a case had been hospitalized with an SSTI as primary diagnosis. In sensitivity analysis, we included secondary diagnoses in the definition of SSTI (i.e. SSTIs not primary cause of hospitalization).

Those who had not been hospitalized with an SSTI (primary or secondary diagnosis) served as controls.

### Genotyping

Details about the genotyping in UK Biobank and HUNT are described in the online supplement.

### Genome-wide association analyses

#### UK Biobank

Genome-wide association analysis was performed in SAIGE (version 0.35.8.3) using a linear mixed model which accounts for cryptic relatedness and imbalance in the proportion of cases and controls.^11^ We included birthyear, sex, genotype chip, and the first six ancestry-informative principal components as covariates. We used SAIGE with same settings to analyze the X-chromosome, coding males as diploid. Variants with minor allele frequency (MAF) > 0.5% were included in the analyses, and dosages were used for imputed variants.

#### HUNT

Genome-wide association tests were carried out by use of SAIGE (version 0.29.4) on autosomal chromosomes,^11^ while BOLT-LMM (version 2.3.4) was used in the analysis of the X-chromosome, coding males as diploid.^12^ Age, sex, genotype batch, and the five first ancestry-informative principal components were included as covariates. Variants with MAF > 0.5% were included in the analyses, and dosages were used for imputed variants.

#### Meta-analysis

We carried out meta-analysis using METAL (version 2011–03–25), with the use of effect size estimates and standard errors as weights, and adjusting for residual population stratification and relatedness through genomic control correction.^13^ A total of 9,211,777 single-nucleotide polymorphisms (SNPs) that were present in both cohorts were included in the meta-analysis.

Associations with p-value <1e–6 and p-value <5e–8 were considered genome-wide suggestive and significant, respectively. Suggestive associations in the discovery cohort were considered replicated in HUNT if the beta coefficient was in the same direction and the p-value was <0.05 divided by the number of independent suggestive loci tested.

#### Gene-based analysis

The summary statistics from the meta-analysis were used in gene-based tests computed by MAGMA (Multi-marker Analysis of GenoMic Annotation) on the FUMA platform by use of the 1000G Phase3 EUR reference panel and default settings.^14,15^ The SNPs were annotated to

19,518 protein coding genes, with p-value threshold for genome-wide statistical significance of 0.05/19,518 = 2.56e–6.

#### Time-to-event analyses

Validation of the results from the case-control analyses were carried out using a prospective design in the HUNT Study. For these analyses, we included cases of SSTI hospitalizations as main diagnosis that occurred after HUNT participation, and subjects were censored when they moved out of the county or at death. We evaluated the cumulative incidence of SSTI by risk genotype and conducted stratified analyses by age and sex.

#### Gene expression

We evaluated the gene expression profiles of genetic loci that were suggestive in the discovery cohort and replicated in the replication cohort. Through the eQTL Catalogue, we accessed gene expression results from 15 published datasets (primarily of immune cells) and from GTEx v8 (general tissues).^16,17^ This allowed us to evaluate single-tissue expression quantitative trait loci and tissue-specific gene expression and regulation.

#### Linkage disequilibrium score regression

LD score regression was used to estimate genetic correlation with traits, conditions and lifestyle factors.^18^ A panel of 1,293,150 well-imputed SNPs that passed quality control was used as reference,^18^ with pre-calculated LD scores for each SNP on European populations from the 1000 Genomes project. Of these SNPs, 1,156,848 common (MAF > 1%) SNPs were available from the meta-analysis of SSTIs. Genetic correlation was assessed between SSTIs and 597 traits, conditions and lifestyle factors registered in the UK Biobank by use of available GWAS summary statistics.

#### Phenome-wide association analysis

Suggestive genetic variants from the discovery cohort that were replicated in the replication cohort were evaluated in phenome-wide association analysis on the Open Targets platform (genetics.opentargets.org), accessing variant-trait-associations from UK Biobank and the GWAS Catalog (accessed July 10^th^, 2020).^19^ Statistically significant associations after correction for multiple testing (conservatively assuming independence between traits) were presented.

#### Mendelian randomization analyses

Leveraging the random allocation of alleles at conception, and that genotype is unchanged over the lifetime, one may use genetic variants as instruments for exposures of interest, thus estimating an association between exposure and outcome that is less susceptible to confounding and reverse causation than traditional epidemiological analyses.^7^ Two-sample MR analyses were conducted separately for the UK Biobank and HUNT Study cohorts. Genetic variants to serve as instruments for the six cardiometabolic exposures of interest – body mass index (BMI), type 2 diabetes mellitus, low-density lipoprotein (LDL) cholesterol, systolic blood pressure, lifetime smoking, and sedentary lifestyle – were extracted from relevant published GWASs (Supplementary Table 1). Only independent SNPs (R^2^< 0.001) with p-value < 5e-8 in these GWASs were included. The TwoSampleMR R package (version 0.5.0)^20^ was used to carry out inverse-variance weighted MR analyses, considered as the main analyses, along with a set of sensitivity analyses using methods that make different assumption about horizontal pleiotropy, including simple median, weighted median and MR Egger.

#### Software

Genome-wide association analyses, meta-analyses, and MR analyses were carried out using R (versions 3.4.4 and 3.6.2), and time-to-event analyses were carried out using Stata MP (version 16.0; College Station, TX, USA).

#### Ethical approval

The Regional Committee for Medical Research, Health Region IV, in Norway (REK) has approved the HUNT study, and this project is regulated in conjunction with The Norwegian Social Science Data Services (NSD). The UK Biobank study has ethical approval from the North West Multi-centre Research Ethics Committee (MREC). Approval for individual projects is covered by the Research Tissue Bank (RTB).

## RESULTS

### Study population

In both the UK Biobank and HUNT, cases, compared with controls, were at baseline older, had higher BMI and systolic blood pressure, and were more likely to be male, ever-smoker and self-reported diabetic (Table 1). In HUNT, cases were more likely to have a sedentary lifestyle, but this information was unavailable to us for cases and controls in UK Biobank.

**Table 1.**
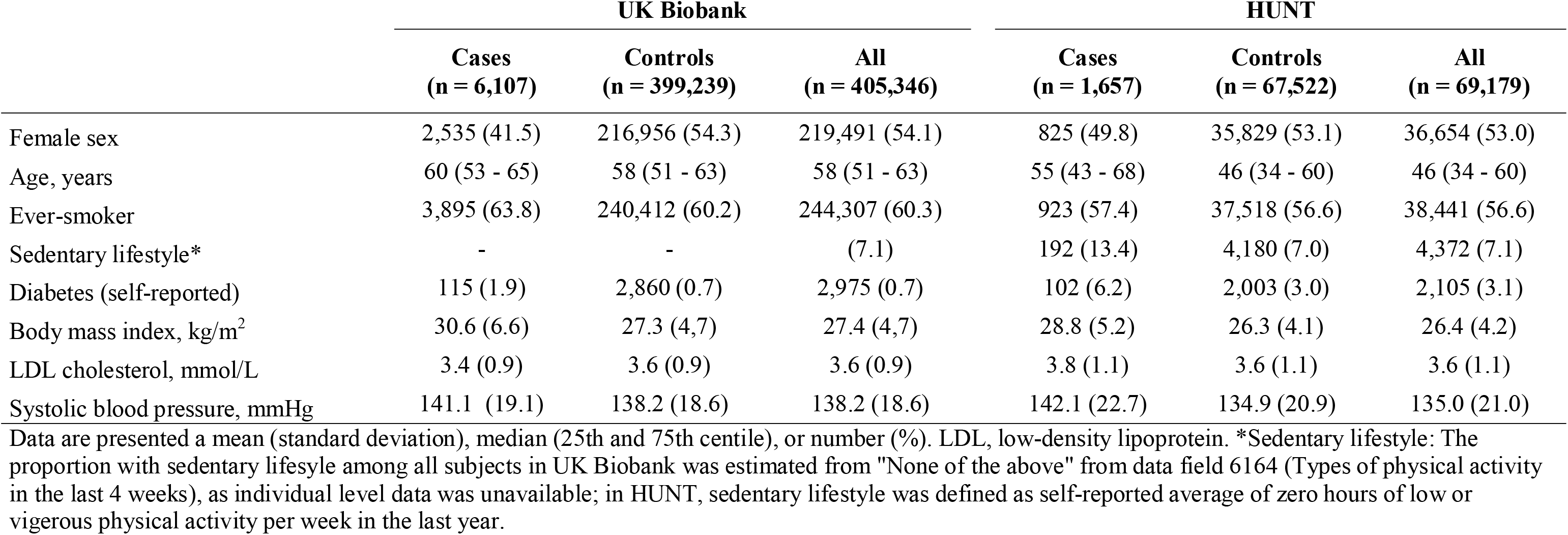
Background characteristics at entry in the UK Biobank and the HUNT Study.

|  | UK Biobank |  |  | HUNT |  |  |
| --- | --- | --- | --- | --- | --- | --- |
|  | Cases<br>(n = 6,107) | Controls<br>(n = 399,239) | All<br>(n = 405,346) | Cases<br>(n = 1,657) | Controls<br>(n = 67,522) | All<br>(n = 69,179) |
| Female sex | 2,535 (41.5) | 216,956 (54.3) | 219,491 (54.1) | 825 (49.8) | 35,829 (53.1) | 36,654 (53.0) |
| Age, years | 60 (53 - 65) | 58 (51 - 63) | 58 (51 - 63) | 55 (43 - 68) | 46 (34 - 60) | 46 (34 - 60) |
| Ever-smoker | 3,895 (63.8) | 240,412 (60.2) | 244,307 (60.3) | 923 (57.4) | 37,518 (56.6) | 38,441 (56.6) |
| Sedentary lifestyle* | - | - | (7.1) | 192 (13.4) | 4,180 (7.0) | 4,372 (7.1) |
| Diabetes (self-reported) | 115 (1.9) | 2,860 (0.7) | 2,975 (0.7) | 102 (6.2) | 2,003 (3.0) | 2,105 (3.1) |
| Body mass index, kg/m<sup>2</sup> | 30.6 (6.6) | 27.3 (4.7) | 27.4 (4.7) | 28.8 (5.2) | 26.3 (4.1) | 26.4 (4.2) |
| LDL cholesterol, mmol/L | 3.4 (0.9) | 3.6 (0.9) | 3.6 (0.9) | 3.8 (1.1) | 3.6 (1.1) | 3.6 (1.1) |
| Systolic blood pressure, mmHg | 141.1 (19.1) | 138.2 (18.6) | 138.2 (18.6) | 142.1 (22.7) | 134.9 (20.9) | 135.0 (21.0) |

Data are presented a mean (standard deviation), median (25th and 75th centile), or number (%). LDL, low-density lipoprotein. \*Sedentary lifestyle: The proportion with sedentary lifesyle among all subjects in UK Biobank was estimated from "None of the above" from data field 6164 (Types of physical activity in the last 4 weeks), as individual level data was unavailable; in HUNT, sedentary lifestyle was defined as self-reported average of zero hours of low or vigerous physical activity per week in the last year.

### Genome-wide association analyses

The main genome-wide association analysis included 6,107 cases and 399,239 controls from the discovery cohort (UK Biobank), and 1,657 cases and 67,522 controls from the replication cohort (HUNT). The association analyses in UK Biobank yielded seven independent loci that were at least genome-wide suggestive, of which two were genome-wide significant (Table 2 and Manhattan plot in Supplementary Figure 1). Corrected for multiple testing (p-value < 5e2–/7 = 7.1e–3), one of the seven suggestive loci replicated in HUNT: lead SNP rs3749748 in the *LINC01184* gene on chromosome 5 (Manhattan plot presented in Supplementary Figure 2). Each copy of the minor allele (T) of rs3749748 (MAF 0.25 and 0.23 in UK Biobank and HUNT, respectively) was associated with an odds ratio (OR) of SSTIs of 1.19 (95% confidence interval [CI] 1.14 – 1.24, p-value = 7.6e–16) in UK Biobank and 1.15 (95% CI 1.06 – 1.25, p-value = 6.3e–4) in HUNT.

**Table 2.** Genetic variants with p-value < 1e–6 in the discovery cohort on risk of skin and soft tissue infection

| Variant name | Chr | Pos | Closest gene | EA/OA | Discovery (UK Biobank) |  |  | Replication (HUNT) |  |  | Meta-analysis |  |
| --- | --- | --- | --- | --- | --- | --- | --- | --- | --- | --- | --- | --- |
|  |  |  |  |  | EAF | OR (95% CI) | p-value | EAF | OR (95% CI) | p-value | OR (95% CI) | p-value |
| rs72989928 | 2 | 210,196,618 | MAP2 | G/T | 0.017 | 0.69 (0.60 - 0.79) | 3.5E-07 | 0.014 | 0.95 (0.68 - 1.33) | 7.7E-01 | 0.72 (0.63 - 0.83) | 2.0E-06 |
| rs62267025 | 3 | 87,726,132 | AC108749.1 | C/T | 0.012 | 1.60 (1.33 - 1.92) | 6.0E-07 | 0.010 | 0.92 (0.63 - 1.35) | 6.6E-01 | 1.44 (1.22 - 1.70) | 2.0E-05 |
| rs150468829 | 5 | 7,081,850 | LINC02196 | A/G | 0.009 | 1.67 (1.36 - 2.05) | 9.7E-07 | 0.009 | 0.98 (0.67 - 1.42) | 9.0E-01 | 1.47 (1.23 - 1.77) | 2.7E-05 |
| **rs3749748** | **5** | **127,350,549** | **LINC01184** | **T/C** | **0.248** | **1.19 (1.14 - 1.24)** | **7.6E-16** | **0.231** | **1.15 (1.06 - 1.25)** | **6.3E-04** | **1.18 (1.14 - 1.23)** | **4.4E-18** |
| rs115740542 | 6 | 26,123,502 | H2BC4 | C/T | 0.075 | 1.23 (1.14 - 1.31) | 7.8E-09 | 0.091 | 1.01 (0.90 - 1.14) | 8.4E-01 | 1.17 (1.10 - 1.24) | 4.2E-07 |
| rs78625038 | 16 | 81,402,279 | GAN | CT/C | 0.006 | 1.98 (1.53 - 2.56) | 2.2E-07 | 0.006 | 1.56 (1.00 - 2.41) | 4.9E-02 | 1.86 (1.48 - 2.32) | 5.9E-08 |
| rs5910356 | X | 117,606,177 | WDR44 | T/C | 0.058 | 0.84 (0.79 - 0.90) | 5.6E-07 | 0.055 | 1.00 (1.00 - 1.00) | 5.9E-01 | 1.00 (1.00 - 1.00) | 7.3E-01 |

Suggestive variants (p-value <1e-6) in the discovery cohort that replicate in the HUNT cohort (p-value < 7.1e-3 and beta coefficient in the same direction) are presented in bold. Chr, chromosome; CI, confidence interval; EA, effect allele; EAF, effect allele frequency; OA, other allele; OR, odds ratio; Pos, chromosome position.

In the meta-analysis of 7,764 cases and 466,761 controls, only the locus in *LINC01184* reached genome-wide significance (Figure 1), while two additional loci were close to genome-wide significance: rs2007361 in *PSMA1* chromosome 11 (additive effect of minor allele OR 0.91, 95% CI 0.88 – 0.94, p-value = 5.1e–8) and rs78625038 in *GAN* on chromosome 16 (additive effect of minor allele OR 1.86, 95% CI 1.48 – 2.32, p-value = 5.9e–8) (regional plots in Supplementary Figures 3 and 4). The genomic inflation factor in UK Biobank (λ = 1.01) did not indicate inflation of the results (quantile-quantile plots in Supplementary Figures 5–7). Sensitivity analyses that included secondary diagnosis of SSTIs as cases (n = 8,652 in UK Biobank and n = 1,900 in HUNT; same number of controls as in the main analyses) yielded very similar, albeit slightly attenuated results (Supplementary Table 2).

**Figure 1.**
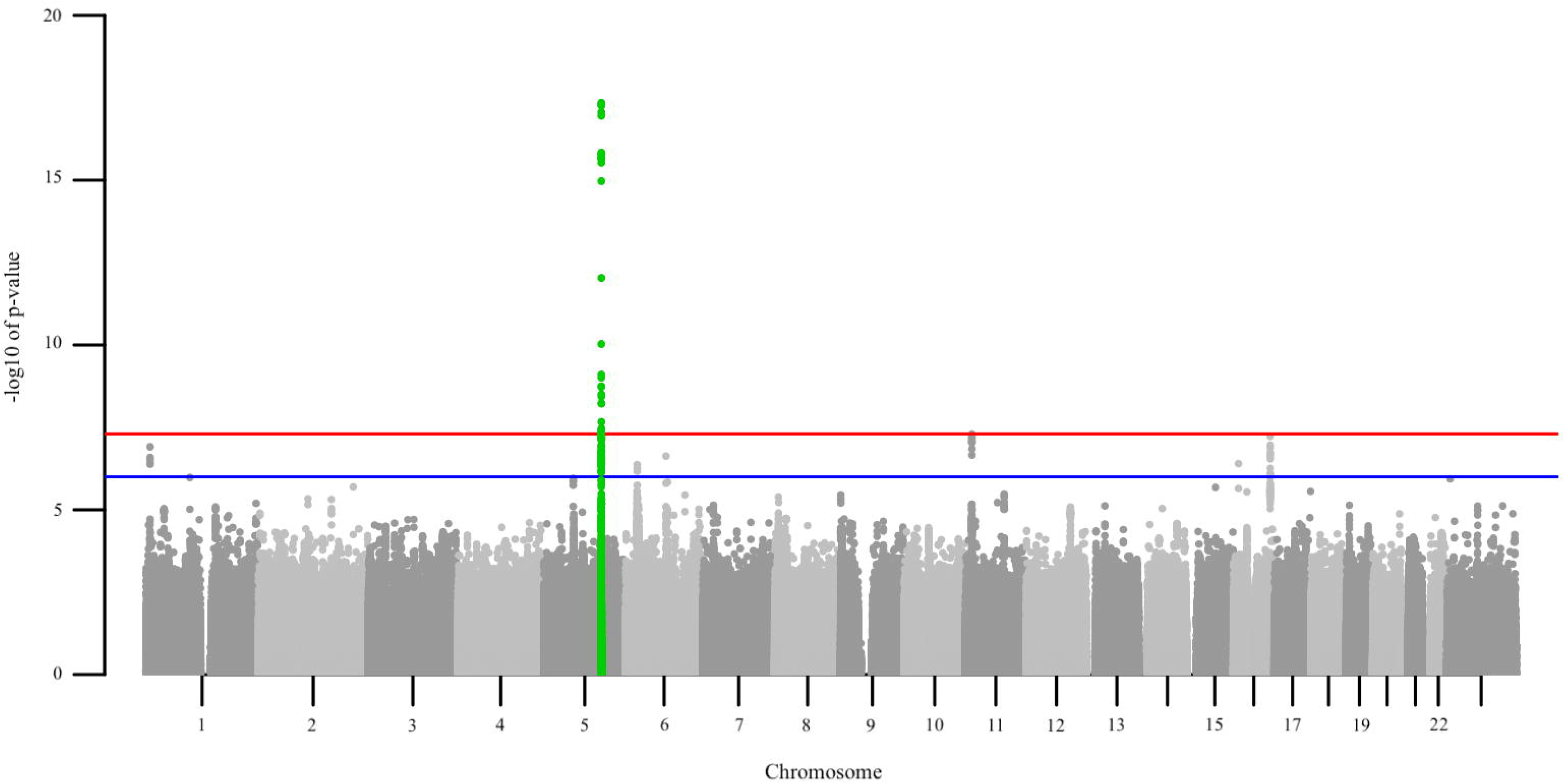
Manhattan plot of results for the meta-analysis.

*Legend:*Axes display the -log10 transformed p-value by chromosomal position. The blue line indicates genome-wide suggestive associations (p-value <1e–6) and the red line genome-wide significant associations (p-value <5e–8). Genome-wide significant loci (+/– 500kb of lead variant) are highlighted in green.

The region that showed the most robust association with SSTI risk was at 127.4 Mb on chromosome 5. While the lead SNP, rs3749748, is closest to *LINC01184*, several SNPs in high LD with this SNP reside in the *SLC12A2* gene (Figure 2).

**Figure 2.**
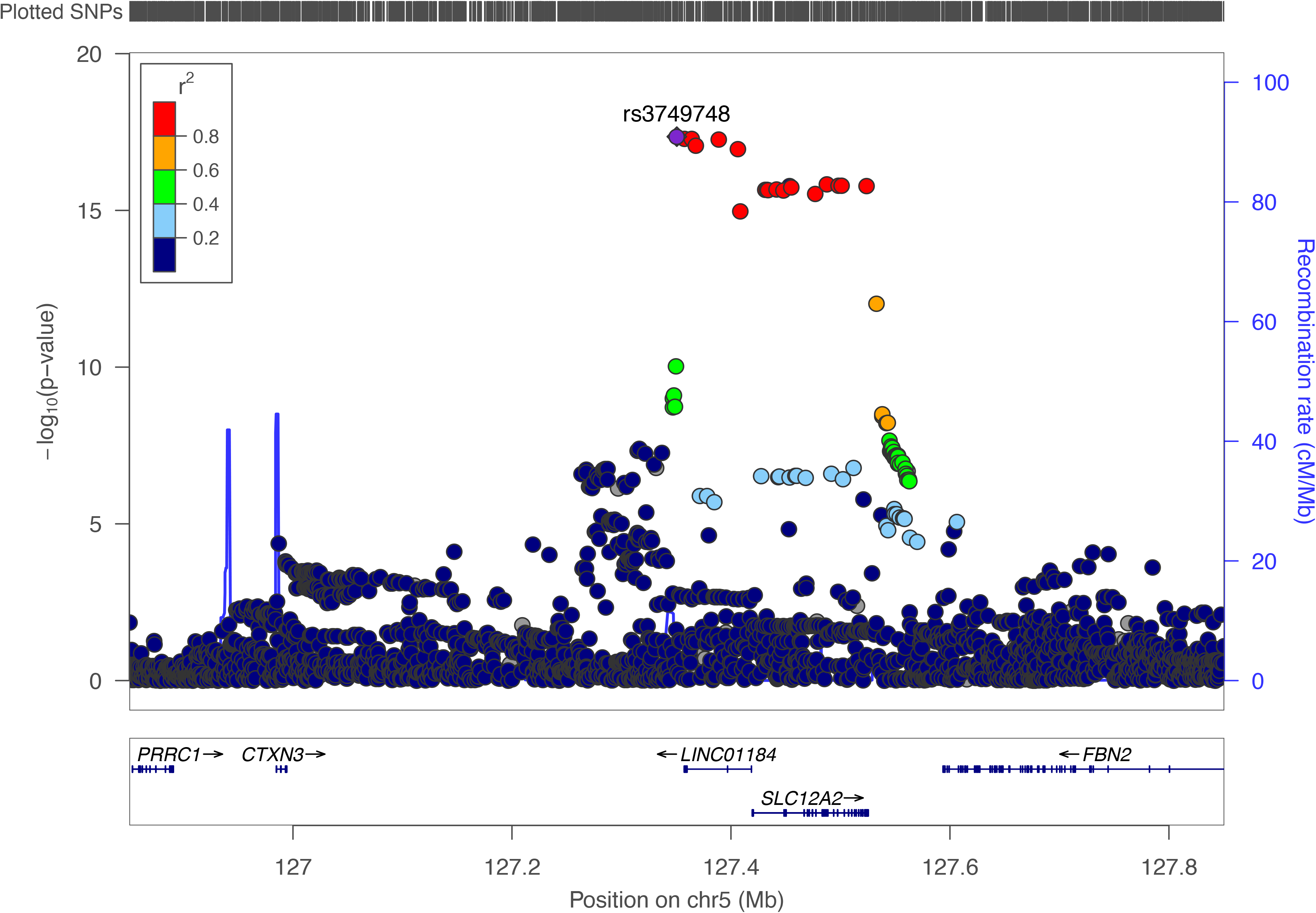
Regional plot of association results of the discovery stage genome-wide significant locus that was replicated.

*Legend* Associations between genetic variants and skin and soft tissue infection from the meta-analysis are plotted by position (x-axis) and -log10 transformed p-values (left y-axis).rs3749748 served as sentinel variant, while the remaining variants are color coded in terms of the linkage disequilibrium (r2) to the sentinel variant. Estimated recombination rates are plotted as light blue lines (right y-axis). The European population from 1000 Genomes Project, November 2014 release, was used as reference, on genome build hg19.

In the gene-based association tests, four genes reached genome-wide significance (Supplementary Figure 8): In addition to the three mentioned loci identified through the SNP-based genome-wide meta-analysis (*LINC01184/SLC12A2, PSMA1* and *GAN*), *IL6R* was associated with SSTI susceptibility.

### Time-to-event analyses

rs3749748 in *LINC01184/SLC12A2* was the only suggestive variant that replicated in HUNT in the genome-wide analyses, and was carried forward to further analyses in the independent HUNT cohort. For these prospective analyses, 69,277 subjects were included, of which 1,496 (2.2%) were hospitalized with an SSTI. In time-to-event analysis stratified by rs3749748 genotype and adjusted for age, sex and five ancestry-informative principal components, the hazard ratio for SSTI hospitalization was 1.17 (95% CI 1.06 – 1.31, p-value = 0.003) and 1.32 (95% CI 1.07 – 1.63, p-value = 0.009) for the CT and TT genotype, respectively, compared with the CC genotype (Supplementary Figure 9). The harmful effect of the CT/TT genotype seemed to exert itself in the young and middle-aged adults, but did not play an important role in risk of SSTI after roughly 70 years of age (Figure 3, left pane). Risk of SSTI was higher in men compared with women, and the CT/TT genotyped had the same effect in both sexes (Figure 3, right pane).

**Figure 3.**
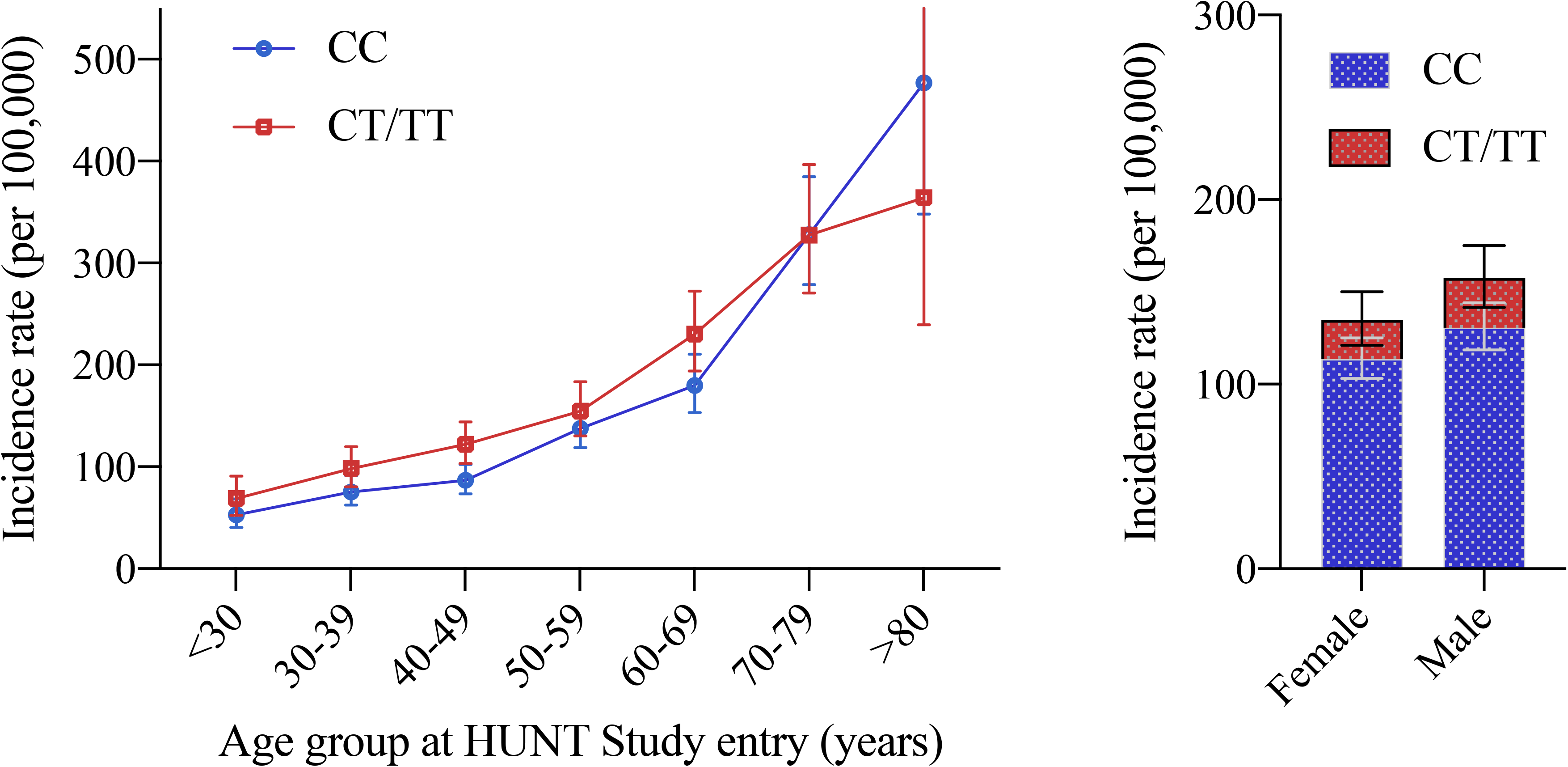
Skin and soft tissue infection incidence rate by age group, sex and rs7949748 genotype.

*Legend*Incidence rates per 100,000 of skin and soft tissue infection hospitalization in HUNT. Blue and red lines/columns represent the incidence for the wildtype (CC) and CT or TT rs7949748 genotype, respectively. Vertical lines represent the 95% confidence intervals.

*Left pane* Stratified by 10-year age groups at study entry. *Right pane:* Stratified by sex.

### Gene expression

In 42 partly overlapping tissues from 15 published datasets evaluated, *SLC12A2* was most highly expressed in skin, while *LINC01184* demonstated highest expression in lymphoblastoid cells (Figure 4). Of the general tissues evaluated in GTEx, the highest median expression of *SLC12A2* was observed in skin, minor salivary glands and the spinal cord, and for *LINC01884* in the cerebellar hemispheres, spinal cord and peripheral nerves (Supplementary Figure 10).

**Figure 4.**
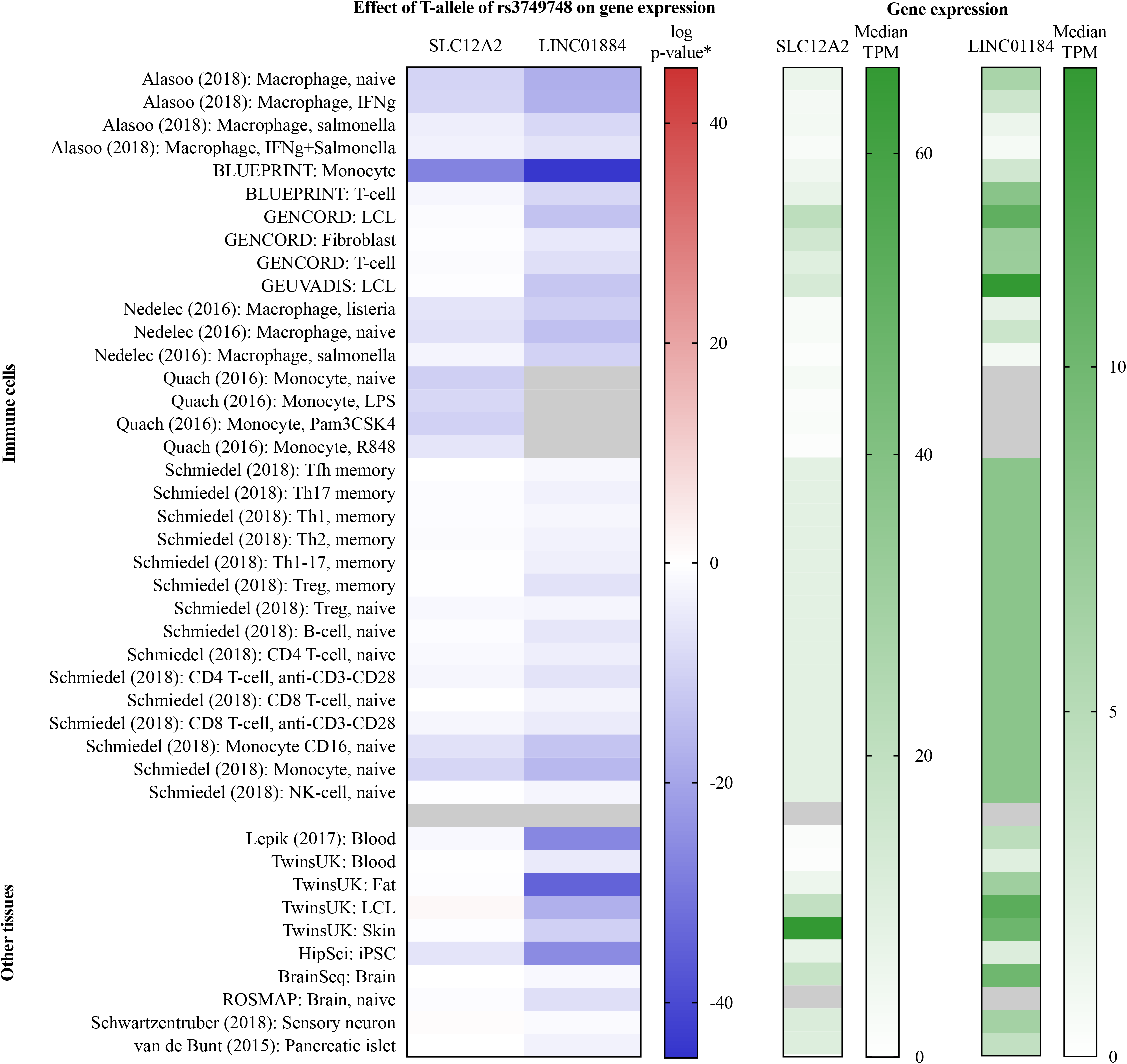
Gene expression of *SLC12A2* and *LINC01884* in 15 published studies.

*Legend* Gene expression of *SLC12A2* and *LINC01884* in the 15 published studies available in the eQTL Catalogue.^16^ Rows are ordered by study and tissue type (immune cells and other tissues). Median TPM by tissue from the RNA sequencing quantification is presented in the right pane, where darker green colors signify greater expression. The effect of the T-allele of rs3749748 on gene expression is presented to the left. *The p-value of the effect was log transformed and given the same sign as the beta coefficient. Thus, blue and red colors signify reduced and increased expression, respectively. Fields without data are colored gray. IFNg, interferon gamma; iPSC, induced pluripotent stem cell; LCL, lymphoblastoid cell line; LPS, lipopolysaccharide; NK, natural killer; Pam3CSK4, Pam3-Cys-Ser-Lys4; resiquimod; Tfh, T follicular helper cell; Th1, T-helper 1 cell; Th2, T-helper 2 cell; Th17, T-helper 17 cell; TPM, transcripts per million; Treg, regulatory T cell.

Presence of the minor allele (T) of rs3749748 was associated with a marked reduced *SLC12A2*-expression primarily in monocytes and macrophages (Figure 4), but was not associated with changed expression in the general tissues evaluated in GTEx (Supplementary Figure 10). *LINC01884*-expression was reduced by the T-allele of rs3749748 in most immune cell lines and general tissues, including monocytes, macrophages, fat, skin, and muscle (Figure 4 and Supplementary Figure 10).

### Linkage disequilibrium score regression

When compared with reported traits, conditions and lifestyle factors in the UK Biobank, SSTIs were the most strongly correlated with cardiometabolic risk factors and self-reported pain. The genetic correlation between SSTIs and the top 15 individual traits, conditions and lifestyle factors are presented in Figure 5, while the complete set of correlations are presented in Supplementary Data 1.

**Figure 5.**
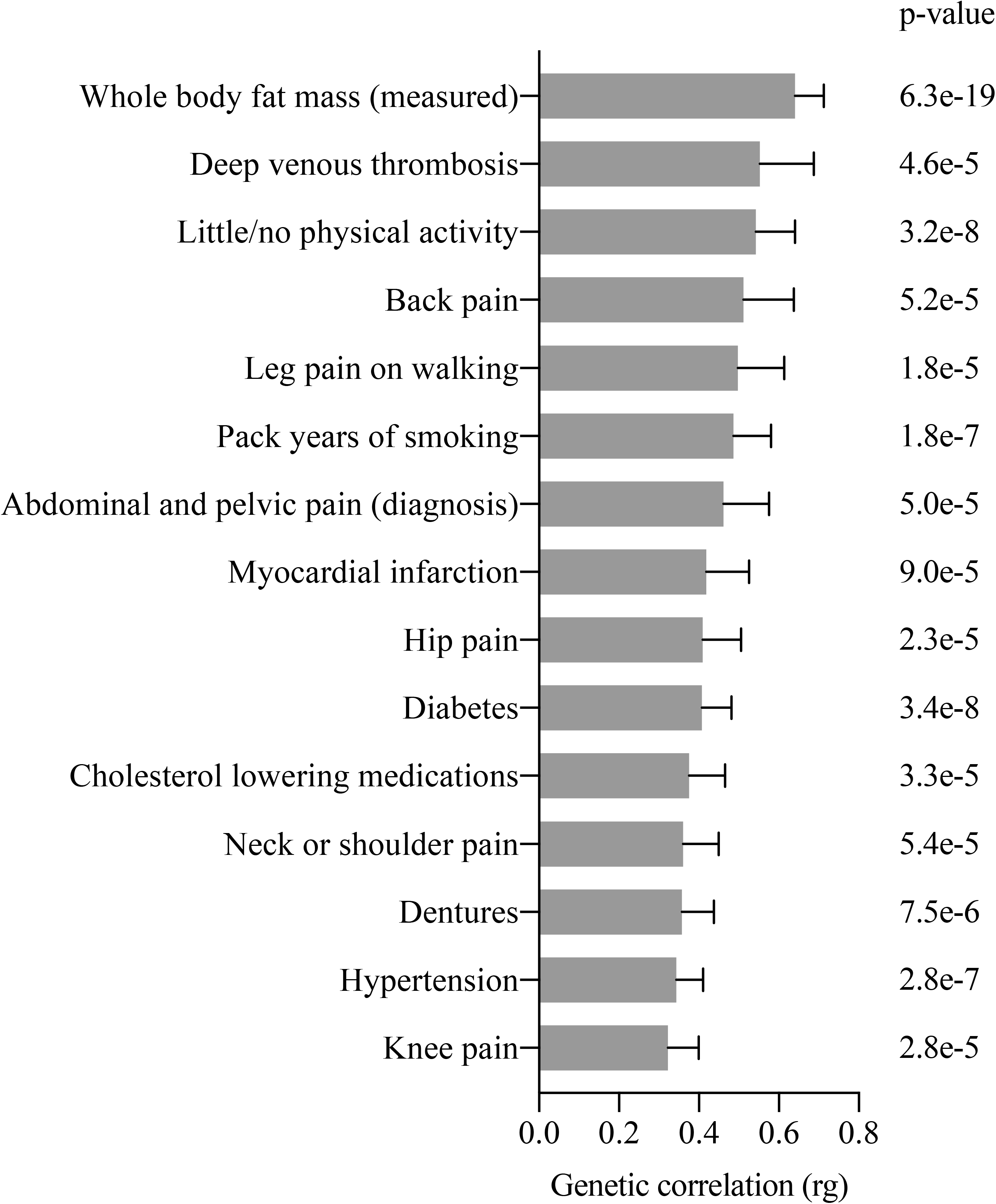
Genetic correlation between skin and soft tissue infection and 15 conditions, traits and lifestyle factors.

*Legend* The 15 distinct conditions, traits and lifestyle factors in the UK Biobank most strongly correlated with skin and soft tissue infection (using data from the meta-analysis). Genetic correlation (rg) presented along the x-axis.

### Phenome-wide association analysis

Phenome-wide association analysis of rs3749748 identified 45 traits or diseases in the UK Biobank and GWAS Catalogue that were statistically significant at p-value < 5e-5 (Supplementary Figure 11). Of note, presence of the T-allele of rs3749748 was associated with body composition such as reduced impedance (lowest p-value = 1.4e–122) and fat percentage (lowest p-value = 1.6e–34), and increased fat-free mass (lowest p-value = 2.2e–47) and whole body water mass (lowest p-value = 3.6e–21); immune and blood cells such as reduced red cell distribution width (lowest p-value = 4.0e–265), immature reticulocyte fraction (lowest p-value = 7.1e–19), and lymphocyte percentage (lowest p-value = 1.8e–11), and increased neutrophil percentage (lowest p-value = 2.01–e1); and other traits and diseases such as increased risk of varicose veins (lowest p-value = 1.7e–20).

### Mendelian randomization analyses

For every standard deviation increase in genetically-predicted BMI, the OR for SSTIs was 1.86(95% CI 1.61 – 2.15, p-value = 1.1e–17) and 1.68 (95% CI 1.29 – 2.19, p-value = 1.4e–4) in UK Biobank and HUNT, respectively (Figure 6). In UK Biobank, every standard deviation increase in genetically-predicted lifetime smoking score was associated with an OR for SSTIs of 2.51 (95% CI 1.75 – 3.61, p-value = 6.4e–7), and in HUNT, OR 2.61 (95% CI 1.31 – 5.17, p-value = 6.1e–3). High systolic blood pressure was found to increase the risk of SSTIs in UK Biobank, and a similar effect estimate – but with less precision – was observed in HUNT. The MR sensitivity analyses supported the findings from the IVW analyses, including no signs of bias due to pleiotropy in the MR Egger analyses of BMI and smoking (Supplementary Table 3).

**Figure 6.**
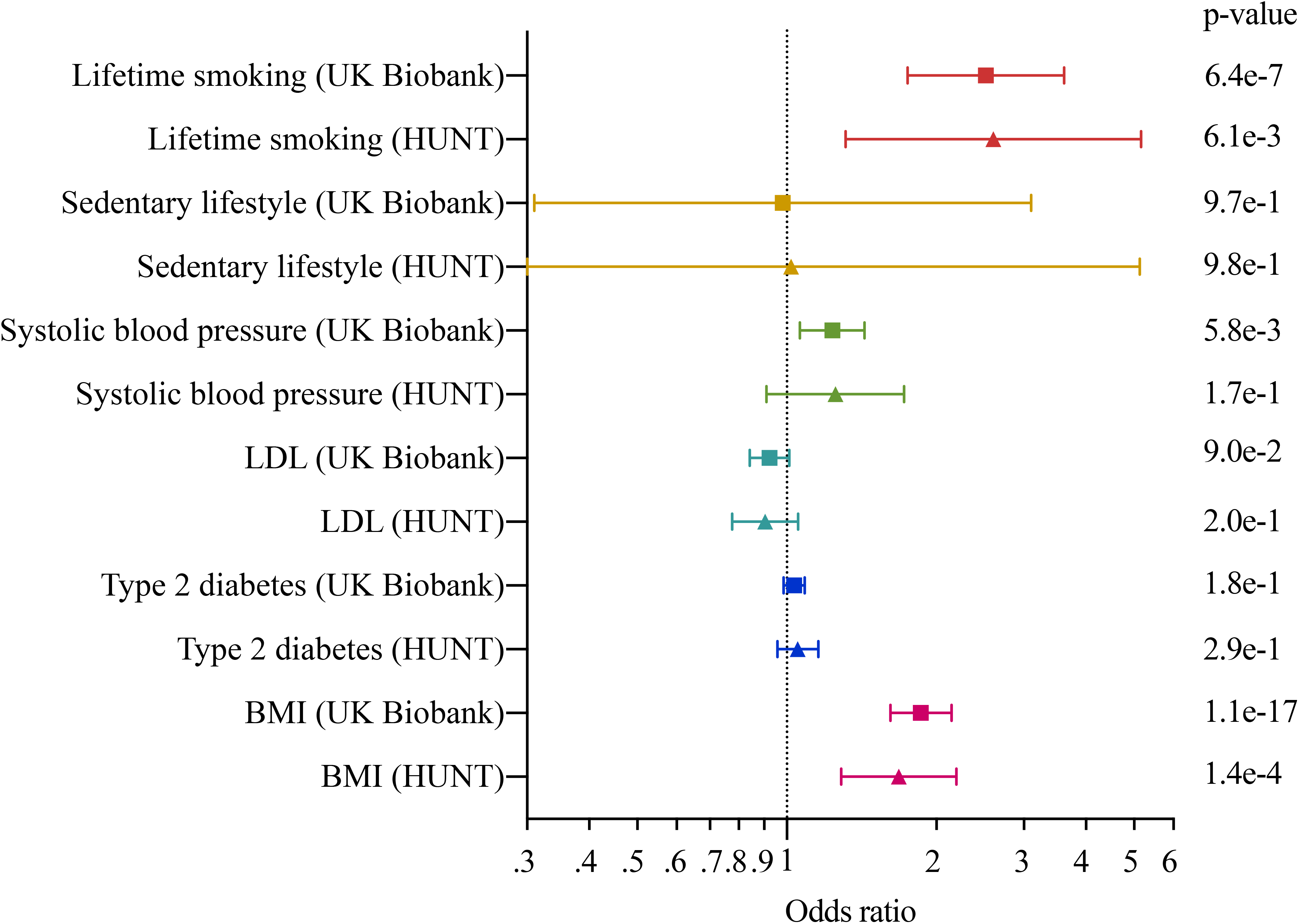
Mendelian randomization analyses of cardiometabolic risk factors on risk of skin and soft tissue infection.

*Legend* Forest plot of the two-sample inverse-variance weighted Mendelian randomization analyses of cardiometabolic risk factors identified as genetically correlated with skin and soft tissue infection. Each risk factor was evaluated separately for UK Biobank and HUNT, and the corresponding risk factors were grouped by color. The x-axis represents the increased odds ratio per standard deviation increase of the genetically predicted risk factor (per unit increase in log odds ratio for genetically proxied type 2 diabetes mellitus liability).

## DISCUSSION

In this first GWAS on SSTI susceptibility we identified a novel risk locus in *LINC01184/SLC12A2* in UK Biobank that was replicated in the independent HUNT Study. Through meta-analysis and gene-based tests, we identified three additional genes associated with SSTI susceptibility; *PSMA1, GAN*, and *IL6R*. Genetically determined higher BMI and lifetime smoking was causally associated with increased risk of SSTIs.

The genetic locus that was by far the most strongly associated with SSTI susceptibility was in *LINC01184/SLC12A2. LINC01184*, or Long Intergenic Non-Protein Coding RNA, is part of the lncRNA class of genes that does not encode for proteins, but have still been found to modulate inflammation and infection risk.^21,22^ *SLC12A2* encodes for the protein NKCC1 which regulates transport of chloride, potassium and sodium across cell membranes, and is key in modulating ion movement across the epithelium, chloride homeostasis, volume of cells, macrophage activation, and anti-microbial activity.^23,24^

Through access to published gene expression studies we observed that across several independent studies, the minor allele (T) of the lead SNP in *LINC01184/SLC12A2*, rs3749748, was particularly associated with reduced gene expression of *SLC12A2* and *LINC01184* in monocytes and macrophages. Macrophages are the most abundant resident immune cell type in the skin, and are critical for the resistance against invading microbes and tissue repair.^25^ The population of skin macrophages are composed of embryonically derived macrophages that self-renew, along with monocyte-derived macrophages.^26^

NKCC1 – along with its kidney-specific isoform NKCC2 – is inhibited by the commonly used loop-diuretic bumetanide.^23^ An experimental study found that macrophages pretreated with bumetanide showed a reduced inflammatory response to bacterial ligands.^27^ Thus, assuming that reduced *SLC12A2* expression in macrophages following variation in rs3749748 leads to reduced functioning of NKCC1, one may hypothesize that bumetanide administration may increase the risk of SSTIs. However, SSTIs have not been identified as adverse events of bumetanide use.^28^ Apart from a small observational study of lower limb cellulitis patients that observed longer hospital stay among users of diuretics (which may be due to common causes such as edema), there are to our knowledge no studies that have reported an association between the use of bumetanide and risk of SSTIs. rs3749748 is associated with a host of other traits, as demonstrated in the phenome-wide association analysis. Many of these traits are linked to cell-volume regulation, which is a key role of NKCC1.^23^ While it is unclear whether any of these traits may themselves be causally linked to SSTI susceptibility, cell-volume regulation may be. By changing the intracellular volume of cells, mechanical barrier function may be regulated and affect immune cell and bacterial migration.^24^

Two loci that almost reached genome-wide significance in the meta-analysis and that were genome-wide significant in the gene-based tests are located in the genes *PSMA1* and *GAN*. The former encodes for a component of the 20S core proteasome complex and is involved in several pathways, including the generation of MHC class-I presented antigenic peptides necessary for the immune response.^29^ *GAN* creates a cytoskeletal component that plays an important role in neurofilament architecture, mediated through ubiquitination and proteasomal degradation.^30^ Finally, gene-based tests identified *IL6R* as associated with SSTIs; a gene that is important in regulation of the immune response, and genetic variation of its ligand, *IL6*, has been linked to SSTI susceptibility.^31^ Interestingly, tocilizumab, a monoclonal antibody against the IL6 receptor, has been linked to increased risk of SSTIs in patients with rheumatoid arthritis.^32^

Cardiometabolic risk factors have been identified as potential causes of SSTIs.^1,5,6^ The links between these traits and SSTIs were reflected in our LD score regression. While LD score regression and traditional epidemiological observations can inform on correlation, they do not necessarily reflect causal effects. Our two-sample MR analyses of six cardiometabolic risk factors identified increasing BMI and lifetime smoking as likely causal risk factors of SSTIs. These effects were consistent across the different MR-methods used, and in the two independent samples, which increases our confidence in these results.^33^ Our finding of BMI as a risk factor for SSTIs align with two recent MR studies,^5,6^ but ours is the first MR-analysis to demonstrate that smoking causally increases the risk of SSTIs.

This study has several strengths, but also some limitations. This is the first GWAS published on SSTIs to date, with a large number of cases and controls. We were able to identify a novel locus – *LINC01184/SLC12A2* – robustly associated with SSTIs in a discovery cohort and replicate the finding in an independent replication cohort. A limitation of our study is that we did not have the power to identify more than one genome-wide significant locus, and we thus encourage replication with meta-analysis in independent cohorts. We only evaluated subjects of European ancestry, and the validity of our findings to other populations is unclear. Of note, while the MAF of rs3749748 in North-Western European populations is around 23%, it is only 4% in African-American populations.^34^ It is therefore important to evaluate populations of different ancestries than the one currently considered. We conducted a range of in-silico analyses, which allowed for identification of monocytes and macrophages as cells of particular interest, and we found strong support for a causal detrimental effect of increasing
BMI and smoking on risk of SSTIs.

In conclusion, we have identified genetic variation in *LINC01184/SLC12A2* to be strongly associated with SSTI susceptibility, and NKCC1, encoded for by *SLC12A2*, is an area of future study in terms of modulation of risk. Interventions to reduce smoking, overweight and obesity in the population will likely reduce the disease burden of SSTIs.

## Data Availability

Data to the UK Biobank and HUNT is available through application. Summary statistics from the genetic association analyses will be made available at the time of journal publication.

## ACKNOWLEDGEMENTS

The Nord-Trøndelag Health Study (The HUNT Study) is a collaboration between HUNT Research Centre (Faculty of Medicine and Health Sciences, Norwegian University of Science and Technology NTNU), Nord-Trøndelag County Council, Central Norway Regional Health Authority, and the Norwegian Institute of Public Health. The authors declare no conflicts of interest.

We gratefully acknowledge all the studies and databases that made GWAS summary data available: ADIPOGen (Adiponectin genetics consortium), C4D (Coronary Artery Disease Genetics Consortium), CARDIoGRAM (Coronary ARtery DIsease Genome wide Replication and Meta-analysis), CKDGen (Chronic Kidney Disease Genetics consortium), dbGAP (database of Genotypes and Phenotypes), DIAGRAM (DIAbetes Genetics Replication And Meta-analysis), ENIGMA (Enhancing Neuro Imaging Genetics through Meta Analysis), EAGLE (EArly Genetics & Lifecourse Epidemiology Eczema Consortium, excluding 23andMe), EGG (Early Growth Genetics Consortium), GABRIEL (A Multidisciplinary Study to Identify the Genetic and Environmental Causes of Asthma in the European Community), GCAN (Genetic Consortium for Anorexia Nervosa), GEFOS (GEnetic Factors for OSteoporosis Consortium), GIANT (Genetic Investigation of ANthropometric Traits), GIS (Genetics of Iron Status consortium), GLGC (Global Lipids Genetics Consortium), GPC (Genetics of Personality Consortium), GUGC (Global Urate and Gout consortium), HaemGen (haemotological and platelet traits genetics consortium), HRgene (Heart Rate consortium), IIBDGC (International Inflammatory Bowel Disease Genetics Consortium), ILCCO (International Lung Cancer Consortium), IMSGC (International Multiple Sclerosis Genetic Consortium), MAGIC (Meta-Analyses of Glucose and Insulin-related traits Consortium), MESA (Multi-Ethnic Study of Atherosclerosis), PGC (Psychiatric Genomics Consortium), Project MinE consortium, ReproGen (Reproductive Genetics Consortium), SSGAC (Social Science Genetics Association Consortium) and TAG (Tobacco and Genetics Consortium), TRICL (Transdisciplinary Research in Cancer of the Lung consortium), UK Biobank.

We gratefully acknowledge the contributions of Alkes Price (the systemic lupus erythematosus GWAS and primary biliary cirrhosis GWAS) and Johannes Kettunen (lipids metabolites GWAS).

This research has been conducted using the UK Biobank Resource under Application Number ‘40135’

